# A consolidated and geolocated facility list in Senegal from triangulating secondary data

**DOI:** 10.1101/2023.05.22.23290283

**Authors:** Daouda M Gueye, Alioune Badara Ly, Babacar Gueye, Papa Ibrahima Ndour, Nancy Fullman, Patrick Y Liu, Khadim Mbaye, Aliou Diallo, Ibrahima Diatta, Saly Amos Diatta, Mouhamadou Moustapha Mane, Gloria Ikilezi, Moussa Sarr

## Abstract

Having a geolocated list of all facilities in a country – a “master facility list” (MFL) – can provide critical inputs for health program planning and implementation. To the best of our knowledge, Senegal has never had a centralized MFL, though many data sources currently exist within the broader Senegalese data landscape that could be leveraged and consolidated into a single database – a critical first step toward building a full MFL. We collated 12,965 facility observations from 16 separate datasets and lists in Senegal, and applied matching algorithms, manual checking and revisions as needed, and verification processes to identify unique facilities and triangulate corresponding GPS coordinates. Our resulting consolidated facility list has a total of 4,685 facilities, with 2,423 having at least one set of GPS coordinates. Developing approaches to leverage existing data toward future MFL establishment can help bridge data demands and inform more targeted approaches for completing a full facility census based on areas and facility types with the lowest coverage. Going forward, it is crucial to ensure routine updates of current facility lists, and to strengthen government-led mechanisms around such data collection demands and the need for timely data for health decision-making.

## Background and Summary

Having comprehensive, routinely updated data on how many health facilities exist – and where they are – is critical for health agency planning and programming operations. From determining investments for new or modified service provision and how to optimally reach underserved communities to supporting efficient medical supply chain and delivery logistics to facility providers,^1–3^ numerous components of health system functioning and performance at least benefit from – if not rely upon – granular health facility data. Furthermore, gaps in such facility information can hinder effective coordination across and within health systems. Beyond important financial and broader resource losses associated with inadequate health system coordination, consequences can quickly escalate when undertaking emergency responses to natural disasters (e.g., 2010 Haiti earthquake^4^) and infectious disease outbreaks.^5^ Nonetheless, many countries have not yet established comprehensive health facility registries, or what is often referred to as a master facility list (MFL),^1,3^ or have done so with corresponding geolocation data (e.g., GPS) and identifiers to support direct linkages across national health information and data systems. This gap between a recognized need – national health facility registries or digitized MFLs – and widespread implementation may not be surprising. After all, the investment case for comprehensive facility data collection and maintenance may often be viewed as less than clear-cut, especially given the high cost and time required to conduct health facility censuses (arguably the ‘gold standard’ for establishing an MFL) and challenges in ensuring full representation across public, private, and informal health sectors with regular updates. Accordingly, efforts to leverage and triangulate existing health facility data sources offer a vital bridge toward building a full national health facility registry or MFL.

Past work demonstrates the utility of triangulating various data sources and supplementary inputs to establish geolocated databases of health facilities.^6–9^ In many ways, Maina and colleagues initially pioneered this approach by assembling a range of government-established MFLs, facility data portals, reports, and other lists to generate a spatial database of publicly managed health facilities for 50 countries in sub-Saharan Africa.^6^ This spatial database offered many strengths, including its well-documented data synthesis process and standardized outputs; at the same time, its comprehensiveness understandably varied by country. For countries where formal MFLs existed as of 2019, this geospatial database directly reflected the equivalent of a national registry for publicly managed facilities. For Senegal, a country with a reported 3,967 health facilities in 2018,^10^ only 1,347 facilities were included and no *case de santés* (health huts), a key publicly managed facility type which offers basic primary care services at the community level, were expressly listed. Country-specific initiatives have drawn from routine health information systems, such as the DHIS2, and sought to harmonize parallel or duplicate facility lists being maintained by disparate entities with formal centralization and verification processes.^11–13^ Data-focused organizations including GRID3 and Bluesquare have both directly supported such triangulation work streams, and then augmented identified gaps or discrepancies in reported GPS coordinates with primary data collection.^8^ Each of these approaches also have strengths, particularly in terms of jumpstarting infrastructure for updating and adding new health facility data over time; however, they require upfront – and longer-term – financial and political commitments to fully implement. Lastly, global platforms such as Human Data Exchange (HDX) and healthsites.io have sought to provide open-sourced health facility data repositories, combining OpenStreetMap functionalities with volunteer-provided information on health facilities.^14,15^ This open-source data approach has various advantages, especially its potential use cases for a wide set of audiences; at the same time, its comprehensiveness is strongly affected by volunteer engagement and participation. A recent WHO endeavor, the Geolocated Health Facility Data (GHFD) initiative, aims to draw from these various approaches and support the establishment of geolocated MFLs for each of the 194 WHO member states by 2027.^16^ To achieve this ambition, particularly for countries without a formal MFL to date, it is important to document approaches used and lessons learned across different resource settings and data contexts.

Senegal has demonstrated regular use of facility-level information and cultivated strong demand for data use in health service planning; however, to the best of our knowledge, Senegal has never had a centralized MFL or comprehensive database of health facilities with directly linkable geolocated information.^6,7^ Many potential use cases and applications for such a consolidated facility list have already been identified, such as strengthening strategic planning and monitoring of health program activities, optimizing resource deployment and logistics to health facilities, and streamlining health service referral systems. Total facility counts, by health region and/or facility type are routinely updated through the Annual Health Map Monitoring Report (*Rapport Annuel de Suivi de la Carte Sanitaire*), as produced by Cellule de la Carte sanitaire et sociale, de la Santé digitale et de l’Observatoire de la Santé (CSSDOS).^17,18^ Furthermore, facility lists known as facility frames have been updated at various times between 2012 and 2019 for Senegal’s continuous Service Provision Assessment (SPA) series to support survey sampling procedures.^10,19–24^ Myriad facility data are publicly available, as well as supported and updated within the country (e.g., COUS facility survey regional facility lists); however, few efforts have occurred to systematically identify and triangulate this range of secondary data into a consolidated facility list. As highlighted by the WHO GHFD initiative and others,^6,8,16^ this first step of data triangulation against existing sources is a critical for paving pathways toward a full MFL or registry equivalent in the future.

Here we describe the approach and process used to triangulate 16 different secondary data sources to build a consolidated and, where possible, geolocated facility list in Senegal from March 2021 to May 2023. These results can serve as a contemporary foundation toward a future MFL in Senegal, identifying areas or facility types with higher levels of georeferencing and those where further (but more targeted) data collection efforts may be most beneficial.

## Methods

### Overview

Our overall approach involved four main steps: 1) identify and collate available facility lists and facility data with GPS; 2) standardize each facility observation across sources and match facilities found in more than one list to each other, with the aim of generating a consolidated list of unique facility observations through a combination of fuzzy-name and geolocation matching, and then manual matching and revision where needed; 3) where available, assign GPS coordinates to each unique facility observation based on existing sources; and 4) conduct additional verification, including review by regional focal points. From January 31-February 1, 2023, a facility list workshop was co-hosted in Dakar by the Institut de Recherche en Santé de Surveillance Epidémiologique et de Formations (IRESSEF), Centre des Opérations d’Urgence Sanitaire (COUS), and Direction de la Planification, de la Recherche et des Statistiques (DPRS) to garner feedback and next steps toward establishing a MFL or equivalent facility database in Senegal.

Going forward, we refer to the resulting facility list from our triangulation and collation procedures as a consolidated facility list, or CFL. Such a designation is meant to reflect the extensive efforts made to consolidate facility information into a list of unique observations while recognizing that the output should not be classified as a formalized MFL. For this CFL, we focus on four main types of health facilities in the formal health sector in Senegal:^25^ hospitals, health centers, health posts, and health huts (Table 1). Facilities classified as “other” – as reported by original sources or designated as “doctors” or an unspecified clinic – are included resulting in datasets^26,27^ but do not contribute to the total counts for the CFL reported here.

**Table 1.** Summary of main facility types in the formal health system in Senegal: hospitals, health centers, health posts, and health huts.

| Facility type | Summary of key facility functions and attributes |
| --- | --- |
| Hospitals | - Provides range of services, from primary to tertiary care<br>- Within the public system, there are national and regional hospitals |
| Health centers | - Main point of service for secondary care, with each health district having at least one health center |
| Health posts | - Main point of service for primary care, serving as the main point of health system contact with most populations<br>- In more rural areas, provides direct oversight and support for health hut service provision<br>- In more urban areas, may be referred to as clinics |
| Health huts | - Provides a subset of basic primary care services as means to extend service access in communities with lower service availability (e.g., rural, remote populations)<br>- Typically linked to a health post or health center for direct supervision and provision of supplies and equipment |

### Data sources

From March 2021 to May 2022, facility-level data were identified and collated from a combination of publicly available data sources, data initiatives such as the Demographic and Health Survey (DHS) program^10,20–24^ or ESRI,^28^ published facility-level datasets such as those by Maina and colleagues,^6^ and data files shared by Senegalese government entities such as COUS and Agence Nationale de Statistique et de la Démographie (ANSD) (Tables 2-3). Table 2 provides information on each source with linked GPS coordinates, while Table 3 summarizes data inputs without GPS. Originally 13 datasets were identified and collated; an additional three datasets were incorporated after the facility workshop in Dakar, bringing the total input facility lists or datasets to 16 by May 2023.

**Table 2.**
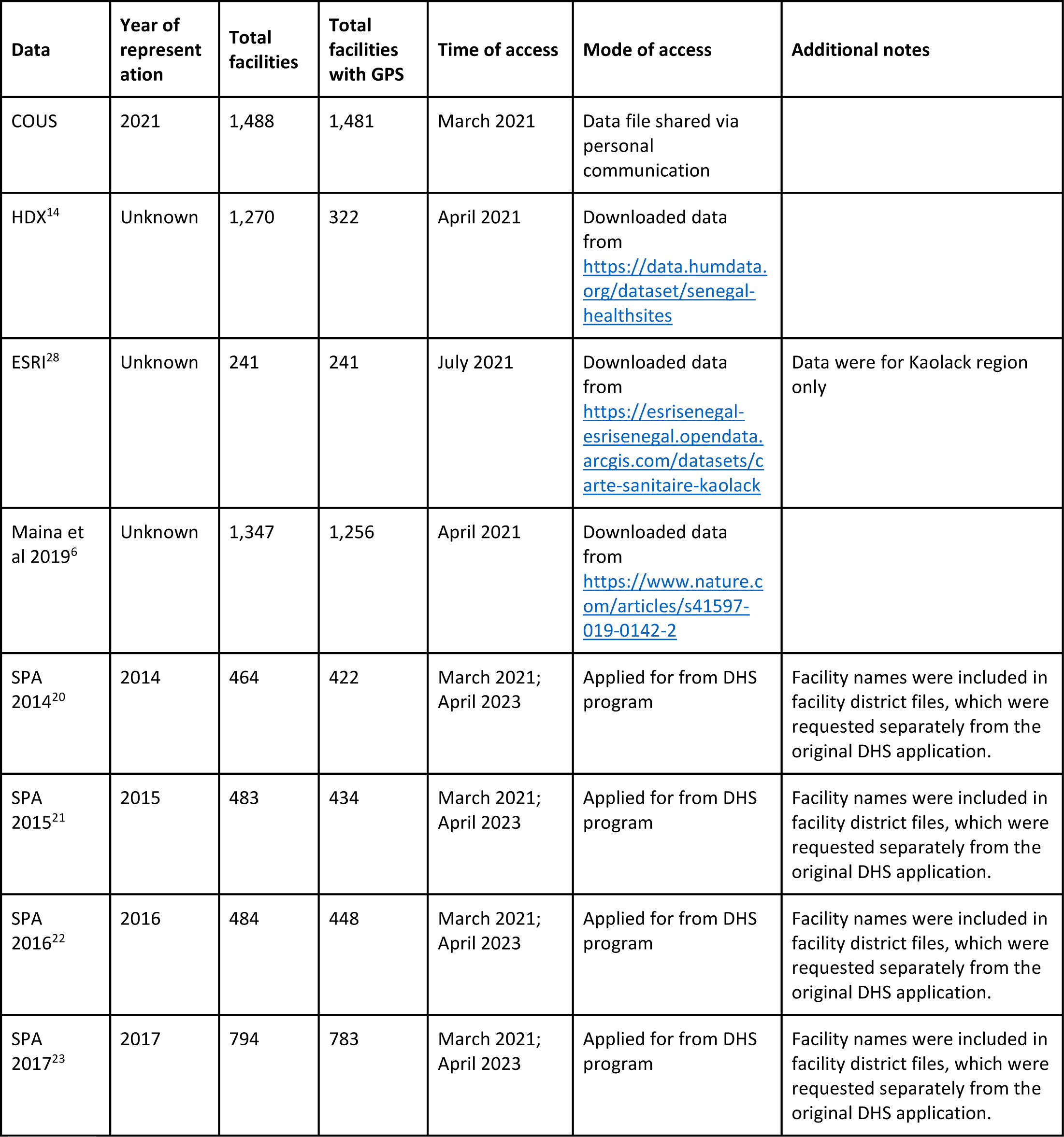
Data sources, with linked GPS, used for triangulation for a consolidated facility list in Senegal. Total facilities reported here do not account for facility duplicates or facilities that were excluded due to being facility types outside of scope for this work (e.g., pharmacies, laboratories); reported values here reflect facility totals as presented in the original data. Total facilities with GPS reported here only include facility types within scope of the CFL: hospitals, health centers, health posts, and health huts. COUS=Centre des Opérations d’Urgence Sanitaire. DHS=Demographic and Health Survey. GPS=Global positioning system. HDX=Human Data Exchange. SPA=Service Provision Assessment.

| Data | Year of representation | Total facilities | Total facilities with GPS | Time of access | Mode of access | Additional notes |
| --- | --- | --- | --- | --- | --- | --- |
| COUS | 2021 | 1,488 | 1,481 | March 2021 | Data file shared via personal communication |  |
| HDX<sup>14</sup> | Unknown | 1,270 | 322 | April 2021 | Downloaded data from https://data.humdata.org/dataset/senegal-healthsites |  |
| ESRI<sup>28</sup> | Unknown | 241 | 241 | July 2021 | Downloaded data from https://esrisenegal-esrisenegal.opendata.arcgis.com/datasets/carte-sanitaire-kaolack | Data were for Kaolack region only |
| Maina et al 2019<sup>6</sup> | Unknown | 1,347 | 1,256 | April 2021 | Downloaded data from https://www.nature.com/articles/s41597-019-0142-2 |  |
| SPA 2014<sup>20</sup> | 2014 | 464 | 422 | March 2021; April 2023 | Applied for from DHS program | Facility names were included in facility district files, which were requested separately from the original DHS application. |
| SPA 2015<sup>21</sup> | 2015 | 483 | 434 | March 2021; April 2023 | Applied for from DHS program | Facility names were included in facility district files, which were requested separately from the original DHS application. |
| SPA 2016<sup>22</sup> | 2016 | 484 | 448 | March 2021; April 2023 | Applied for from DHS program | Facility names were included in facility district files, which were requested separately from the original DHS application. |
| SPA 2017<sup>23</sup> | 2017 | 794 | 783 | March 2021; April 2023 | Applied for from DHS program | Facility names were included in facility district files, which were requested separately from the original DHS application. |

**Table 3.**
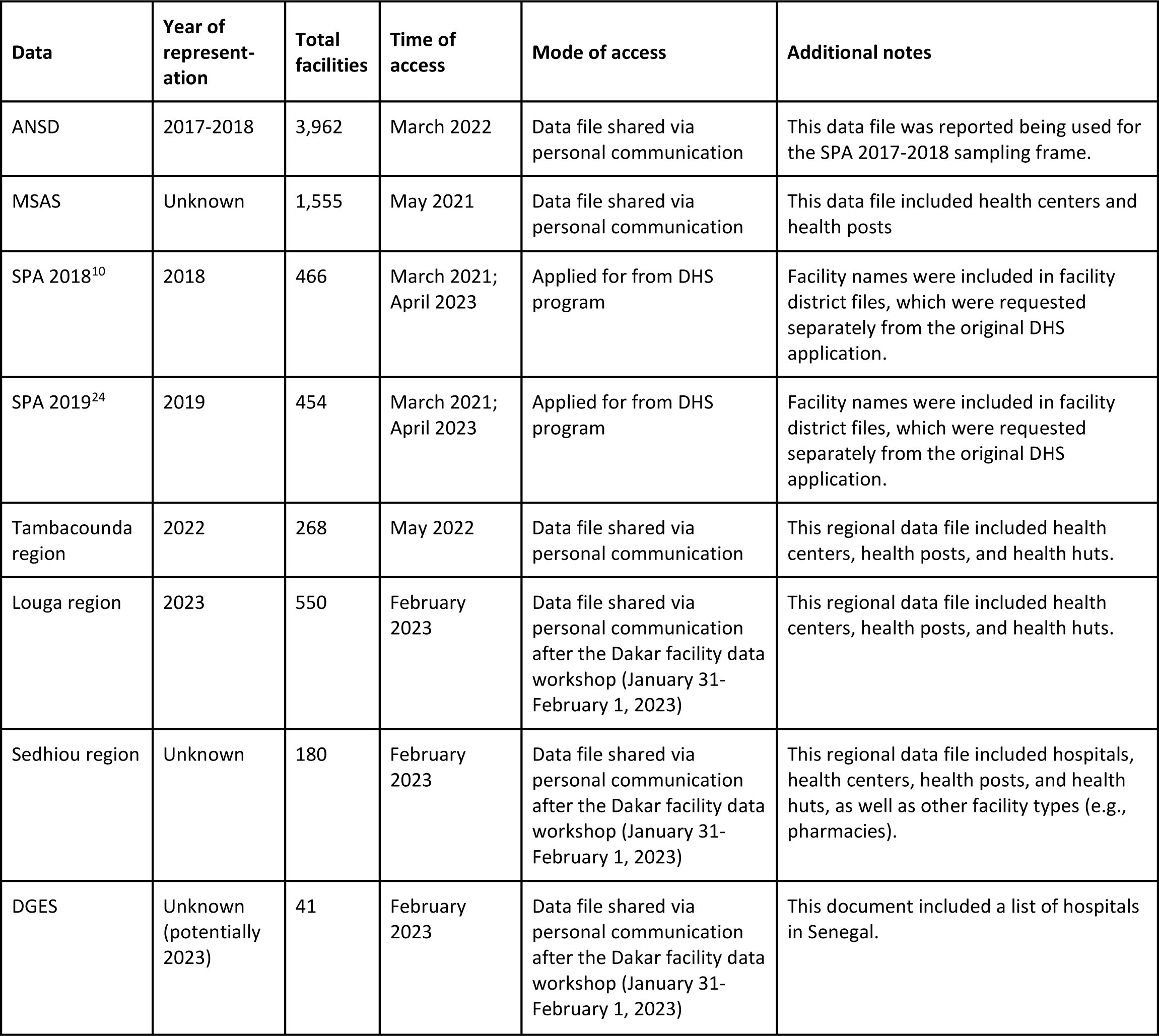
Data sources, without linked GPS, used for triangulation for a consolidated facility list in Senegal. Total facilities reported here do not account for facility duplicates or facilities that were excluded due to being facility types outside of scope for this work (e.g., pharmacies, laboratories); reported values here reflect facility totals as presented in the original data. ANSD=Agence Nationale de la Démographie et de la Statistique. DGES=Direction Générale des Etablissements de Santé. DHS=Demographic and Health Survey. GPS=Global positioning system. MSAS=Ministère de la santé et de l’Action Sociale. SPA=Service Provision Assessment.

| Data | Year of representation | Total facilities | Time of access | Mode of access | Additional notes |
| --- | --- | --- | --- | --- | --- |
| ANSD | 2017-2018 | 3,962 | March 2022 | Data file shared via personal communication | This data file was reported being used for the SPA 2017-2018 sampling frame. |
| MSAS | Unknown | 1,555 | May 2021 | Data file shared via personal communication | This data file included health centers and health posts |
| SPA 2018<sup>10</sup> | 2018 | 466 | March 2021;<br>April 2023 | Applied for from DHS program | Facility names were included in facility district files, which were requested separately from the original DHS application. |
| SPA 2019<sup>24</sup> | 2019 | 454 | March 2021;<br>April 2023 | Applied for from DHS program | Facility names were included in facility district files, which were requested separately from the original DHS application. |
| Tambacounda region | 2022 | 268 | May 2022 | Data file shared via personal communication | This regional data file included health centers, health posts, and health huts. |
| Louga region | 2023 | 550 | February 2023 | Data file shared via personal communication after the Dakar facility data workshop (January 31-February 1, 2023) | This regional data file included health centers, health posts, and health huts. |
| Sedhiou region | Unknown | 180 | February 2023 | Data file shared via personal communication after the Dakar facility data workshop (January 31-February 1, 2023) | This regional data file included hospitals, health centers, health posts, and health huts, as well as other facility types (e.g., pharmacies). |
| DGES | Unknown (potentially 2023) | 41 | February 2023 | Data file shared via personal communication after the Dakar facility data workshop (January 31-February 1, 2023) | This document included a list of hospitals in Senegal. |

### Data processing, matching, and verification steps

Fig. 1 provides an overview of the key steps involved in data processing, facility matching, and verification steps undertaken for this CFL.

**Figure 1.**
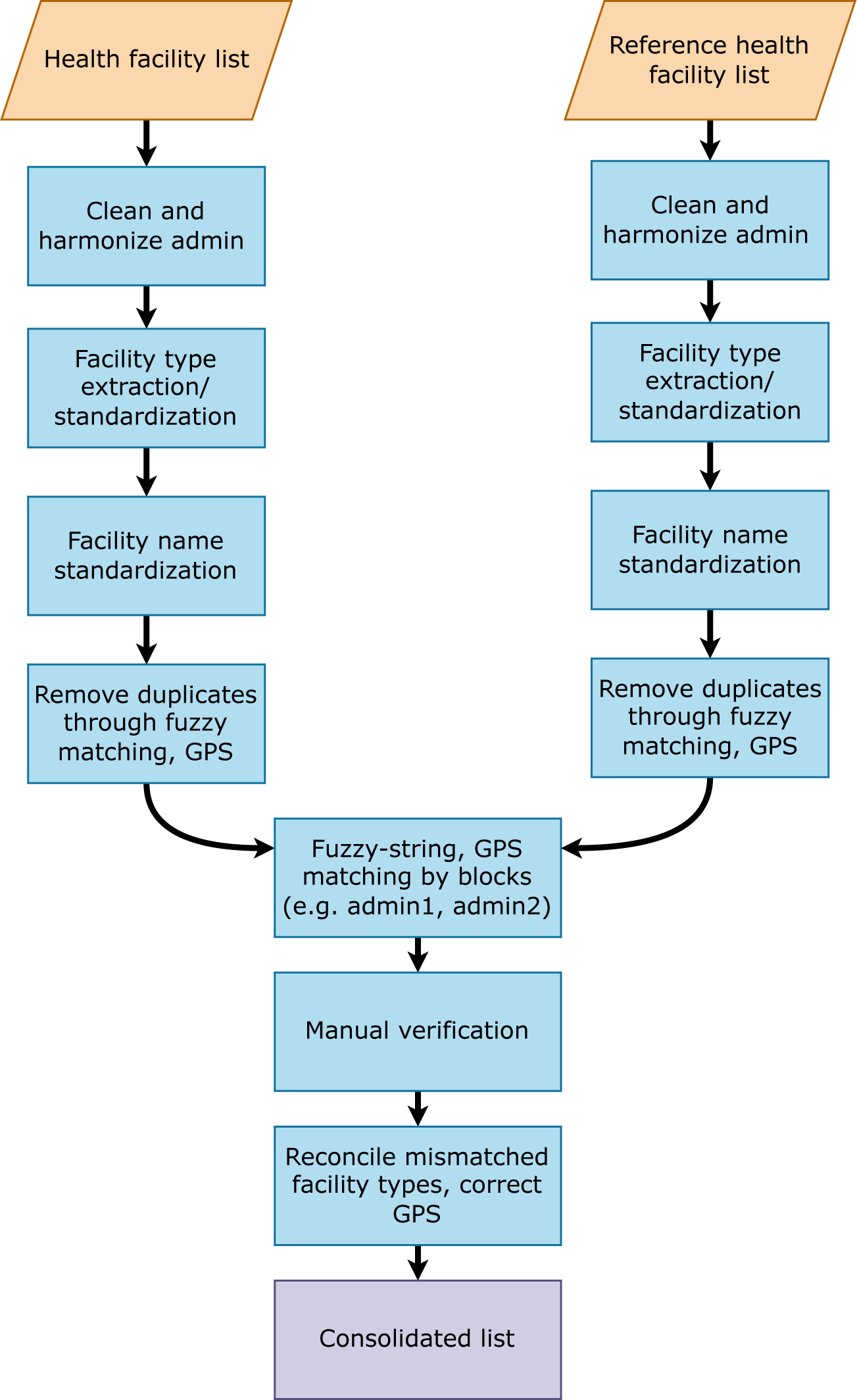
Overview of facility list data processing, matching and verification steps.

For each data source, we extracted and standardized the following facility attributes: facility name, facility type, facility ownership, and first-level administrative unit (region). When not provided in the original dataset, facility type was identified by the facility name string (e.g., “PS Nemataba” or “Nemataba Poste de Santé” in Kolda). Duplicate facilities within sources were removed.

We then combined all facility observations into a pooled database (*n*=12,965 facility observations). Within the pooled, de-duplicated facility database, we used a fuzzy-matching algorithm to group facility names within each region as potential matches. This algorithm was operationalized as a Jaro-Winkler distance,^29^ with a threshold score of 0.2. To optimize the matching process, we first pre-processed the facility name string by removing non-Latin characters, removing facility type strings (e.g., “CS”, “Case de Santé), expanding common acronyms (e.g., “St.” to “Saint”), and standardizing facility numbering (e.g., “Facility iii” to “Facility 3”). Matches that met the fuzzy-string threshold were then manually verified and corrected for any false positive and false negative matches.

The matched lists underwent additional technical and manual verification. For the technical verification, we identified and reviewed all facility matches where the implied distance between observations, as defined by the Haversine distance between GPS points,^30^ exceeded 1 kilometer (km), as well as matches with multiple or conflicting facility types across sources. The entire database was then carefully reviewed to assess the accuracy of matches and identify facilities where additional verification was needed by national and/or regional health system experts in Senegal. While identified verification needs varied, the most common areas for further confirmation included:

● *Same facility name with different facility types in a given region (and often department and health district)* Facilities with the same name but different facility types can be distinct facilities that should be treated as unique observations in a CFL (e.g., CS Fatick and Hôpital de Fatick in the region of Fatick). However, more frequently these facilities were once classified as a lower-level facility that have been subsequently upgraded to a higher level facility type (e.g., a health hut being upgraded to a health post). Accordingly, they are technically the same facility (with the same GPS), and should be treated as a single facility observation for the CFL.
● *Same facility name and type in a given region but different departments and/or health districts*. Facilities with the same name and facility type but are listed under different second-level administrative units usually fall into one of two scenarios. First, they are in fact distinct facilities and a given region has more than one of the same facility type with the same name (e.g., PS Mlomp in the health districts of Oussouye and Thionk Essyl in the region of Ziguinchor); subsequently these facilities should be treated as unique observations in a CFL. Second, differences in second-level administrative unit names may actually reflect different administrative categorizations that both technically comprise the “second-level” geographic groupings after Senegal’s 14 regions: departments (n=45) and health districts (n=77 to 79). Departments are the formal second-level administrative boundaries in Senegal, whereas health districts reflect the geographies at which peripheral health authorities operate the country’s health system structure.^25^ Although we mapped health districts to corresponding departments for 98% facility observations, further reconciliation is needed.
● *Same facility type in a given region (and often department and/or health district) with similar but ‘different enough’ names*. Facilities with similar but ‘different enough’ names usually are not matched via algorithms. When GPS are available for a set of facilities in question, manual matches can be ascertained based on how similar their locations are. However, unless additional triangulation can be done, these facilities are flagged for further verification.

The next step involved assigning available GPS coordinates for each ‘unique’ facility observation. If a singular facility had linked GPS coordinates or a matched group of facilities across sources only had one set GPS associated with them, the corresponding GPS coordinates were used. If multiple GPS coordinates were reported across sources for a given facility group, we applied a preferential algorithm for source-specific GPS: COUS, and if no GPS from COUS, then non-2017 SPA (2016, 2015, 2014), then SPA 2017, then ESRI, then HDX, and then Maina and colleagues. This hierarchy was based on the degree to which source-specific GPS coordinates appeared to vary and prioritizing government-associated sources (COUS and then SPA, which was co-implemented by ANSD) over external datasets. Last, we supplemented GPS assignment with Google Maps for 9 hospitals from DGES, most of which had opened since 2020. No additional geolocation activities occurred.

Three rounds of additional data verification occurred by sharing the CFL and corresponding inquiries to regional focal points for each of Senegal’s 14 medical regions and health system experts. All regions participated in the first round of verification, which occurred from June 2022 to August 2022. After incorporating the initial feedback provided by regional focal points, we sought to conduct follow-up verification efforts to address the remaining flagged facilities. Four regions (Dakar, Diourbel, Fatick, and Thies) provided second-round feedback from November to December 2022. The initial version of this CFL was disseminated by IRESSEF, COUS, and DPRS at a two-day workshop in Dakar, Senegal from January 31-February 1, 2023; the workshop report is available in Supplementary file 1. Nine out of 14 medical regions were in attendance, with Sedhiou and Louga focal points providing additional feedback and facility lists for verification purposes. A representative from the Direction Générale des Etablissements de Santé (DGES) also provided an up-to-date list of hospitals in Senegal; in combination, these three additional data sources were used as a third round of verification.

After data processing and verification procedures, we generated a CFL of 4,685 unique facility observations (Supplementary table 1),^26^ with 2,423 of these facilities having at least one set of linked GPS. Fig 2a shows the distribution of all geolocated facilities in Senegal, while Fig 2b-e reflect these distributions by facility type (hospitals, health centers, health posts, and health huts).

**Figure 2.**
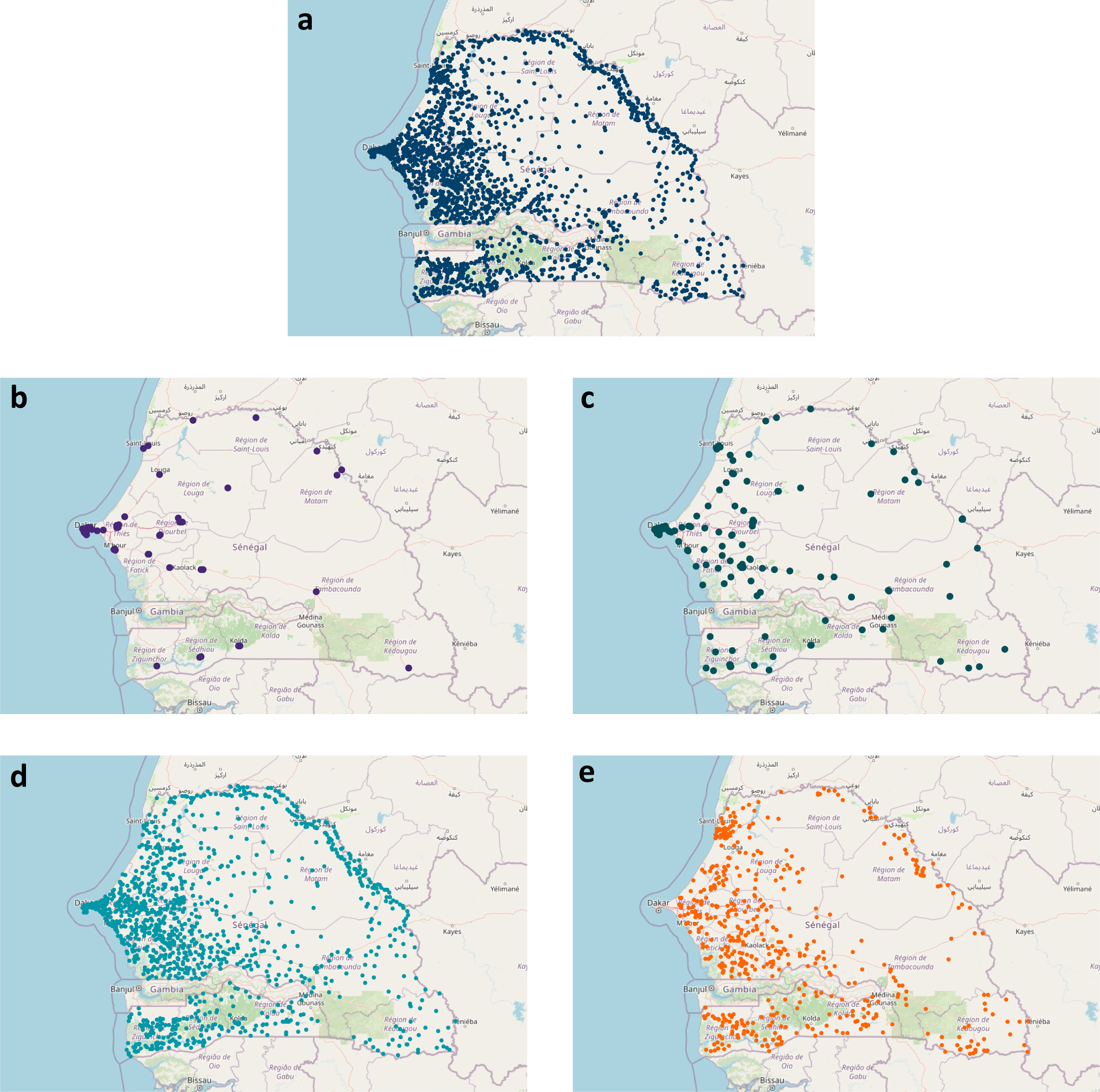
Distribution of geolocated health facilities in Senegal. a All geolocated facilities. b All geolocated hospitals. c All geolocated health centers. d All geolocated health posts. e All geolocated health huts. Facility counts and percentages with GPS, nationally and by region, for all facilities and by facility type can be found in Supplementary table 1.

## Data Records

The CFL and full facility list described here are available publicly and freely at the following repository through both the figshare repository^26,27^ and GitHub: https://github.com/iressef-egh/senegal-cfl The datasets available through figshare will remain in their original form, while the data hosted at GitHub will be updated as further data validation or updates occur. Table 4 details variables in the CFL data file, while Supplementary table 2 lists variables and descriptions in the full facility list dataset, both of which are available in .csv formats; equivalent codebooks in French and English are on GitHub.

**Table 4.** Variable descriptions for the consolidated facility list in Senegal. Variable descriptions for the full facility list are available in Supplementary table 2.

| Consolidated facility list dataset |  |
| --- | --- |
| File name | senegal_consolidated_facilitylist.csv |
| Total observations | 4,685 hospitals, health centers, health posts, and health huts |
| Consolidated facility list variables |  |
| Variable name | Variable description |
| region | Region in Senegal. |
| match_id | Facility id assigned for each unique facility grouping identified in the consolidated facility list. This identifier links the unique observations found in this dataset to the ‘full’ version of this dataset where all facilities for a given match_id have been matched into match groups.<br>*Note:* this id does not correspond with a particular data source or formal health information system (e.g., DHIS2). |
| match_name | This is the processed version of health facility name used for matching, excluding special characters and variations found across sources. Original facility names, as included by source, can be found under “fac_name_orig” in the “senegal_full_facilitylist.xlsx” dataset. |
| group_fac_type | This is the facility type assigned to a unique facility observation. Original facility types may vary by source, of which are listed under “fac_type_orig” in the “senegal_full_facilitylist.xlsx” dataset. Facility types included in this dataset are as follows in French (without special characters), with English translations:<br>- hopital= hospital<br>- centre de esante = health center<br>- poste de esante = health post<br>- case de sante= health hut<br>- autre = other |
| group_latitude | Assigned latitude coordinates. |
| group_longitude | Assigned longitude coordinates. |
| group_gps_source | Source of assigned facility GPS. |
| n_gps | Number of linked sources with GPS for a unique facility observation.<br>*Note:* For a given facility, if multiple GPS coordinates were available and matched precisely, they were not counted as distinct sets of GPS. |
| n_source | Number of linked sources for a unique facility observation. |
| source_list | List of sources for a unique facility observation. |
| data_flagged | Variable whereby “1” indicates outstanding verification needs or follow-up questions. |
| data_notes | Further detail about outstanding verification needs or follow-up questions about the given facility. *All notes are in English at present.* |

The CFL and full facility list data files are meant to be complementary to each other, with the CFL providing a more streamlined dataset with the most internally consistent variables across input data. The full facility list dataset, which is linkable to the CFL with the match_id variable, provides more granular information that can be directly transported to the CFL based on user decisions or preferences. For instance, health facility ownership varies substantially across matched facilities (i.e., 1,895 facilities [40.4%] had different managing authorities or ownership associated within a unique facility group [Supplementary table 3]). Accordingly, using the match_id variable, users can determine which sources they view as more accurate to assign facility ownership.

In terms of geographic characteristics, both datasets include region – the first-level administrative unit for Senegal – and GPS coordinates are rounded off to five decimal points for consistency. All coordinates are reported in decimal degrees format, per the World Geodetic System 1984 (WGS84) coordinate system. Departments (second-level administrative unit) and health districts are provided in the full facility list file, as they are not as consistently available across all facilities. More localized administrative units in Senegal – arrondissements and communes – were not included for most input data sources.

## Technical Validation

### Geolocation of facilities

Nationally, 51.7% of these unique facility observations had at least one set of linked GPS coordinates, though the relative percentage of GPS representation varied by region (Supplementary table 1). By facility type, hospitals and health centers generally had the highest proportion of GPS coverage. For hospitals, 9 out of 14 regions had 100% GPS coverage, while Kaolack was the main exception (with 25% of reported hospitals had GPS); this low coverage may be a function of facility type discrepancies between ESRI Kaolack and other sources. For health centers, 88.3% of facilities had GPS assigned through the triangulation process. Seven regions had 100% of identified health centers as also geolocated while five regions had GPS coverage for health centers at less than 80%: Louga (56.7%), Kaffrine (66.7%), Sedhiou (71.4%), and Diourbel (72.7%). For Louga and Sedhiou, at least some of the lower GPS coverage is likely related to the availability of very recent facility data sources without geolocation (e.g., 2023 for Louga). Across health posts nationally, 67.9% had GPS coverage; such comparatively low coverage was largely skewed by Dakar, where only 32.5% of facilities designated as health posts had GPS coordinates. This is likely associated with the high prevalence of private clinics in Dakar,^31^ for which more granular facility information can be more challenging to access. Outside of Dakar, GPS coverage ranged from 91.9% in Saint-Louis to 72.8% in Tambacounda; it is worth noting that the lower GPS coverage for Tambacounda is likely associated with the inclusion of a non-geolocated regional facility list of health centers, health posts, and health huts for 2022. For health huts, GPS coverage was comparatively low nationwide (26.5%); the exception was Diourbel, where 28 of the 33 identified health huts had at least one set of GPS.

Among facility matches with GPS (*n*=2,423), 1,433 had multiple distinct GPS coordinates linked to unique facility observations (Supplementary table 4). Among facilities with multiple potentially viable GPS, 26.8% had GPS coordinates exceeding a 2 km distance from each other – a potential marker for additional verification (Supplementary table 4).

### Number of health facilities, by type and geographically

In addition to the verification processes described earlier, we identified five sources against which to cross-reference CFL’s total numbers of facilities, nationally and by region (and facility type): reported facility frame numbers as extracted from the SPA 2012-2013, SPA 2017, and SPA 2019 reports,^19,23,24^ and the CSSDOS Health Map reports for 2019 and 2021 (public facilities only).^17,18^ Since the SPA facility frames were used to select facilities ultimately surveyed, these extracted metadata could serve as proxy “MFL” envelope against which to compare the CFL outputs.

At the national level, the CFL had more total facilities that of all other previously published lists (Table 5); however, the Health Map 2021 report only reported on public facilities. There was more variation across previous lists by facility type (Table 5; Supplementary table 5), which likely reflects differences or changes in facility classifications over time.

**Table 5.**
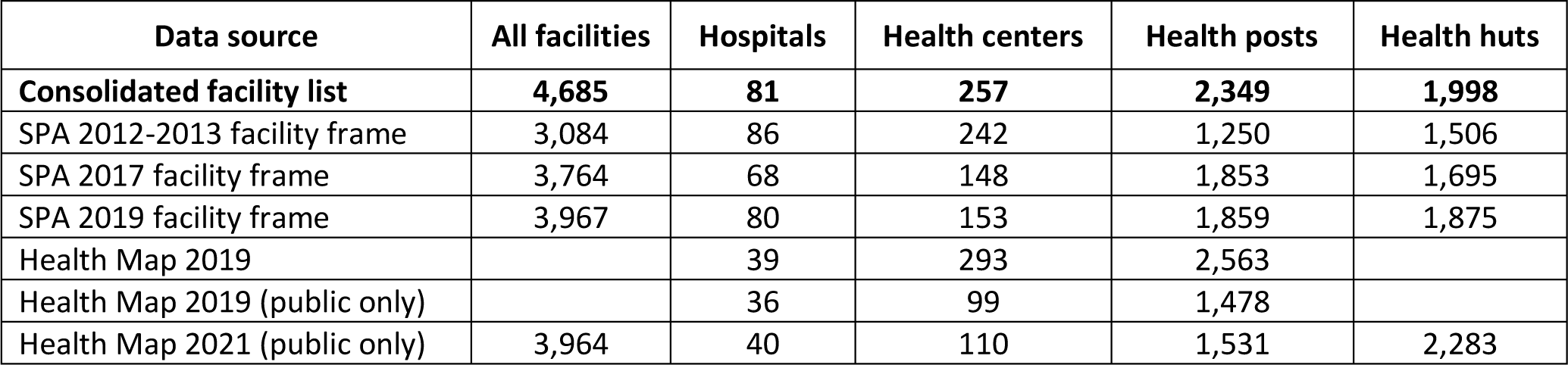
Comparing number of facilities in the consolidated facility list with facility numbers reported in previously published facility lists by facility type at the national level.

Regionally, and then by facility type within regions, greater variation emerged (Supplementary table 5). Such differences likely reflect a combination of data considerations and challenges, including propagation of facility duplicates through published data sources (e.g., not systematically accounting for facility upgrades or changes and thus potentially including the same facility twice under different facility types in a given year) and inconsistent classification across facility types, especially among regions with larger urban centers (and potentially higher prevalence of private facilities). For instance, 486 facilities (10.4% of the total CFL) originally had more than one associated facility type within a unique match across sources (Supplementary table 3). This was particularly prevalent among health centers (27.2% of 257) and health posts (15.2% of 2,349), with the latter being largely due to facilities being upgraded from health huts. For other cases, having multiple original facility types may be related to earlier misclassification or potential errors in original data entry. For instance, many health centers were reported as hospitals by SPA surveys, and 11 facilities classified as health posts in Maina and colleagues – which did not report on health huts – were categorized as health huts by other sources. In rare cases, higher-level facilities were reclassified or reopened as a lower-level facility. For instance, PS Ninefecha in Kedougou was originally a hospital from 2002 to 2013, but was reopened as a health post in 2014.^32,33^ SPA surveys conducted in 2014, 2015, and 2016 classified the facility as a hospital, as did the SPA 2018 survey; the SPA 2017 survey, the COUS survey, and a MSAS facility list all reported Ninefecha as a health post. Of note, 202 of the 486 facilities (41.6%) with more than one associated facility type had their facility type resolved via further triangulation or verification by regional focal points.

### Number of geolocated facilities compared with other published databases

Lastly, we compared the number and distribution of health facilities with GPS from the CFL to those compiled in a spatial database of public facilities by Maina and colleagues (Supplementary table 6).^6^ At the national level, the CFL provides nearly twice as many geolocated facilities (*n*=2,423) as the number of facilities with GPS in Maina and colleagues (*n*=1,256). The inclusion of health huts, with 530 geolocated, accounts for a substantive portion of these differences. Another main contributor was the exclusive focus on public facilities. For instance, in Dakar, the CFL included 797 total facilities, with 328 having GPS; in contrast, only 121 facilities with GPS were included for Dakar in the Maina and colleagues’ database.

## Usage Notes

With these data, we provide a consolidated list of health facilities in Senegal across 16 facility data sources and thus offer an important first step toward building a full MFL in the future. With 52% of facilities with at least one set of GPS coordinates, there is ample need for further prioritizing geolocation efforts if more geospatially targeted health service planning and delivery is to be better supported nationwide. This is particularly important for health huts, which serve as critical connectors between rural communities and higher levels of care. We view this CFL as not only a data product on its own, but also a potential tool that can inform future efforts around where (and which types of facilities) likely have the lowest GPS coverage to date. In looking to the future, an essential next step likely involves augmenting government infrastructure and processes around developing – and routinizing – a MFL.

This CFL has a number of potential applications and use cases in Senegal, of which would only be bolstered by the establishment of a full MFL. At a high level, more targeted and responsive planning could occur, as well as more coordinated public health programming. As highlighted elsewhere,^8,34^ building a geolocated MFL or registry equivalent can provide vital inputs into more geospatially tailored service provision and delivery, such as optimizing community care site placements and microplanning for vaccination services. Another example could involve addressing physical access barriers for antenatal or delivery care; with over 60% of in-facility births occurring at health posts in 2019,^35^ having a more precise understanding of how close – or how far away – maternal health services are could augment models of care and referral networks. By establishing more formalized linkages between a geolocated MFL and health information systems like DHIS2, more granular assessments of facility capacity relative to population need can be performed. For instance, this CFL and eventual MFL in Senegal could inform estimates of COVID-19 vaccination administration capacity, per Africa CDC targets of vaccinating 60-70% of currently eligible populations.^36^ This CFL also can serve as an important input into onward assessments of disease risk (e.g., malaria risk stratification maps^37^), as well as analyses quantifying proximity or access to the nearest facility. In addition, linking estimates of maternal and neonatal mortality relative to facility-level variations and measures of quality could further inform national efforts around meeting Sustainable Development Goal targets for maternal and child health.^38–40^

Beyond the Senegal context, it is possible that other groups – regionally, within sub-Saharan Africa, and/or more globally – could foster cross-country learning opportunities around different approaches to triangulating existing data and implementing more targeted geolocation efforts. This kind of knowledge exchange could facilitate adaptations or innovations in the ways that facility data are routinely collected, verified, and updated for MFLs or registries. Lastly, the confluence of these work streams could support WHO’s GFHD initiative,^16^ including inputs into the GHFD’s global facility database.

In addition to the more discrete obstacles to establishing a MFL (e.g., geolocating the full range of health facility types and managing authorities), it is vital to determine the best ways to routinize MFL data collection, maintenance, and linkages to current health data infrastructure. Establishing formal data governance mechanisms and leadership by governmental department(s) or agencies are likely among the most crucial steps in this process. In addition, parallel collaboration at the national and subnational levels around health facility list verification and updating is essential, particularly in more decentralized health systems and settings where a mixture of data systems operate (e.g., some combination of digital or centrally databased health information and individual or paper-based file systems). This may be particularly relevant for Senegal and potential strategies around integrating a consolidated list and eventual MFL with DHIS2, given the country’s decentralized data processes at regional or health district levels and harmonization with community-level health platforms (e.g., DHIS2 does not directly connect with health huts, which channel paper-based data up to their supervising facilities).^41^

### Limitations and future improvements

The current version of this CFL is subject to several limitations, of which can be considered areas for future improvements. First, this dataset should not be viewed as an MFL, contemporaneously or at a given time in the past. This CFL could not be calibrated against a full facility sampling frame, facility census, or documentation of currently (or ever) operational facilities. While we sought to conduct such benchmarking with the ANSD 2017-2018 dataset, the facility file did not match facility counts by type or region for either the SPA 2017 or SPA 2018 facility frames.^23,24^ As such, the CFL presented here should be viewed as a first step toward establishing a future MFL in Senegal and can help identify key areas where further investment in geolocation may be needed.

Second, it is likely that the private sector remains under-represented in this CFL, both in terms of total facilities and those with GPS. This is at least partly due to the composition of available facility data inputs (i.e., several data inputs included only public facilities), as well as higher missingness or non-reporting of GPS among private facilities in facility surveys. We sought to derive GPS locations based on facility addresses provided through a 2016-2017 assessment of private facilities in Senegal;^31^ however, nearly all of the addresses included insufficient information for Google-based tools to appropriately assign reasonable GPS coordinates. Progress toward a full MFL with GPS will require improving the health data landscape for the private sector, particularly in areas with a higher prevalence of private health service delivery (e.g., Dakar).

Third, this CFL does not systematically track the evolution of facility type changes and/or name changes over time. Since this endeavor sought to identify unique facility observations across datasets, merged facilities that were reported as having been upgraded or changed facility types were assigned their most recent facility type in the CFL. Original facility types, when available, were retained in the ‘full’ version of the CFL.^27^ In terms of name changes, we sought to account for these instances wherever possible; however, they were opportunistic adjustments rather than a systematic undertaking of revisions.

Fourth, the current CFL does not comprehensively capture the opening and closure of facilities over time. Such information would be important to incorporate into future efforts to routinize such facility data. Master facility registries in Malawi^42^ and Ethiopia^43^, as well as prior facility data collected in Ethiopia,^39^ include information on facility opening dates; accordingly, facility cohorts can be constructed over time and support more granular tracking of facility service availability and their contributions to intervention coverage.^39^

Fifth, 347 facilities are still flagged for further review or confirmation. Most of these facilities are either facilities with different facility types with the same name or facilities with the same name associated with different second administrative units; in the absence of GPS to cross-reference a potential match, they are being treated as separate facilities at present. Further engagement with focal points in five regions (Dakar, Diourbel, Fatick, and Thies) from November-December 2022 supported additional reconciliation for at least some flagged facilities, as did the sharing of regional datasets from Louga and Sedhiou after the Dakar-based workshop. In the datasets published alongside this paper,^26,27^ we have sought to clearly identify facilities for which further review should occur.

Sixth, further reconciliation is needed for discordant GPS coordinates across data sources in the CFL. At present, when multiple GPS coordinates are linked to a facility, we use a fairly subjective algorithm to assign GPS. The ‘full’ version of the facility list provides all GPS associated for a given facility,^27^ which could support more sophisticated geospatial testing and verification of discordant GPS coordinates. In addition, facilities with only one source of GPS warrant additional geospatial verification. Such corroboration could be achieved through different approaches, including further triangulation with future health facility surveys; the use of routine health data systems (e.g., prompting health facility managers to input GPS coordinates for flagged facilities via DHIS2), or leveraging existing health campaigns and outreach activities (e.g., piggybacking facility geolocation alongside polio and measles vaccination campaigns^8^).

Seventh, more granular administrative levels, such as arrondissement and communes (i.e., third- and fourth-level administrative units, respectively), have not yet been systematically ascribed to facility-levels observations. Such information was not included in the vast majority of input datasets (i.e., only ESRI Kaolack, the Louga regional list, and the MSAS facility list had them provided), and a number of facilities remained flagged as having discordant health district or department assignments – second-level administrative units – across lists. As GPS coverage is expanded and facility coordinates are confirmed, it may be beneficial to also verify associated arrondissement and commune data. However, given that health districts are considered the lowest level of health sector administration in Senegal,^25^ focusing efforts to verify total facility counts and geolocation by district may be of higher priority.

Last, subjective decisions were made throughout this work, including data availability, inclusion, and processing. While we have sought to provide as much documentation as possible around each step, it is possible that errors or miscoding of information occurred (especially around manual revisions and re-matching). In the ‘full’ version of the facility list,^26^ we provide additional information on decisions made and original facility names and sources so that onward revisions or updates can occur based on the source inputs.

With this CFL, we provide a foundation from which a more comprehensive, geolocated MFL could be developed in Senegal – an important first step toward strengthening data-informed health planning and programs throughout the country.

## Data Availability

The dataset supporting this article is available at the following repository: https://github.com/iressef-egh/senegal-cfl

## Code Availability

Matching algorithms were developed in R,^44^ while manual review and verification occurred using MS Excel (Microsoft, Redmond, USA). Figures were also created in R. All code can be found on GitHub, under ∼/resources/code: https://github.com/iressef-egh/senegal-cfl

## Acknowledgments

We are grateful to all MSAS departments and affiliated agencies, whose overall support and willingness to facilitate data access made this work possible. We especially thank the regional focal points who provided feedback at various points in this work, including attending the 2023 facility list workshop. We thank health facilities and their staff, who gave of their time to complete the health facility surveys used in these datasets. We appreciate the guidance and assistance provided by the DHS program around facility data applications. We acknowledge Audrey Ihler of the Institute for Health Metrics and Evaluation (IHME) for work on geolocating private facilities, as well as IHME team members who gave feedback on this work as part of the Exemplars in Vaccine Delivery project in Senegal. We thank the following individuals from Gates Ventures, for their programmatic support and assistance: Niranjan Bose, David Phillips, Jordan-Tate Thomas, Rowan Hussein, and Ryan Fitzgerald.

## Author contributions

DMG, ABL, PIN, NF, PYL, KM, and AD were involved in identifying, applying for, and/or sharing data. DMG, NF, PYL, and KM were involved in data collation, analysis, and review, with DMG leading regional focal point verification. DMG, NF, and PYL were involved in conceptualizing the database. DMG, NF, PYL, GI, and MS were involved in writing the initial draft of the manuscript. All authors were involved in the revision of the manuscript. All authors read and approved the final version of the manuscript.

## References

1. McFarlane, T. D., Teesdale, S. & Dixon, B. E. Chapter 12 - Facility Registries: Metadata for Where Care Is Delivered. in Health Information Exchange (ed. Dixon, B. E.) 183–201 (Academic Press, 2016). doi:10.1016/B978-0-12-803135-3.00012-8.

2. Noor, A. M., Gikandi, P. W., Hay, S. I., Muga, R. O. & Snow, R. W. Creating spatially defined databases for equitable health service planning in low-income countries: the example of Kenya. Acta Trop 91, 239–251 (2004).

3. World Health Organization (WHO). Master facility list resource package: guidance for countries wanting to strengthen their master facility list: facilitator guide for the MFL training. (WHO, 2019).

4. Rose-Wood, A. et al. Development and use of a master health facility list: Haiti’s experience during the 2010 earthquake response. Global Health: Science and Practice 2, 357–365 (2014).

5. Noor, A. M. et al. Mapping the distribution and risk of epidemics in the WHO African Region/Cartographie de la distribution et des risques d’epidemie dans la Region africaine de l’OMS. Weekly Epidemiological Record 93, 251–257 (2018).

6. Maina, J. et al. A spatial database of health facilities managed by the public health sector in sub Saharan Africa. Sci Data 6, 134 (2019).

7. South, A. et al. A reproducible picture of open access health facility data in Africa and R tools to support improvement. Wellcome Open Res 5, 157 (2020).

8. Geo-referenced Infrastructure and Demographic Data for Development (GRID3). Mapping Health Facilities. https://academiccommons.columbia.edu/doi/10.7916/d8-v5kc-5227/download (2021).

9. Burstein, R. Mapping child health: statistical applications for high-resolution estimation of child mortality and healthcare utilization. (2018).

10. Agence Nationale de la Statistique et de la Démographie (ANSD) & ICF. Senegal: Enquête Continue sur la Prestation des Services de Soins de Santé (ECPSS) 2018. https://dhsprogram.com/publications/publication-SPA32-SPA-Final-Reports.cfm (2020).

11. Darcy, N. et al. Case Study: The Tanzania Health Facility Registry. in Healthcare Policy and Reform: Concepts, Methodologies, Tools, and Applications 339–368 (IGI Global, 2019). doi:10.4018/978-1-5225-6915-2.ch017.

12. Makinde, O. A. et al. Development of a Master Health Facility List in Nigeria. Online J Public Health Inform 6, e184 (2014).

13. Mishra, A. & Sahay, S. Building a master health facility list: innovative Indian experience. BMJ Innovations 7, (2021).

14. Humanitarian Data Exchange (HDX). https://data.humdata.org/.

15. healthsites.io. https://healthsites.io/.

16. World Health Organization (WHO). Geolocated Health Facilities Data initiative. https://www.who.int/data/GIS/GHFD.

17. Cellule de la Carte sanitaire et sociale, de la Santé digitale et de l’Observatoire de la Santé (CSSDOS). Rapport Annuel de Suivi de la Carte Sanitaire 2019. https://www.sante.gouv.sn/sites/default/files/Carte%20sanitaire%20Senegal%20Rapport%20annuel%20de%202019_0.pdf (2020).

18. Cellule de la Carte sanitaire et sociale, de la Santé digitale et de l’Observatoire de la Santé (CSSDOS). Rapport Annuel de Suivi de la Carte Sanitaire 2021. https://www.sante.gouv.sn/sites/default/files/RAPPORT%20ANNUEL%20DE%20SUIVI%20DE%20LA%20CARTE%20SANITAIRE%202021.pdf (2022).

19. Agence Nationale de la Statistique et de la Démographie (ANSD) & ICF. *Sénégal Enquête Continue sur la Prestation des Services de Soins de Santé (ECPSS) 2012-*2013. https://dhsprogram.com/publications/publication-SPA18-SPA-Final-Reports.cfm (2013).

20. Agence Nationale de la Statistique et de la Démographie (ANSD) & ICF. *Sénégal Enquête Continue sur la Prestation des Services de Soins de Santé (ECPSS)* 2014. https://dhsprogram.com/publications/publication-SPA21-SPA-Final-Reports.cfm (2015).

21. Agence Nationale de la Statistique et de la Démographie (ANSD) & ICF. *Senegal Enquête Continue sur la Prestation des Services de Soins de Santé (ECPSS)* 2015. https://dhsprogram.com/publications/publication-SPA25-SPA-Final-Reports.cfm (2016).

22. Agence Nationale de la Statistique et de la Démographie (ANSD), Ministère de la Santé et de l’Action Sociale (MSAS), & ICF. *Sénégal Enquête Continue sur la Prestation des Services de Soins de Santé (ECPSS)* 2016. https://dhsprogram.com/publications/publication-SPA26-SPA-Final-Reports.cfm (2017).

23. Agence Nationale de la Statistique et de la Démographie (ANSD), Ministère de la Santé et de l’Action Sociale (MSAS), & ICF. *Senegal: Enquête Continue sur la Prestation des Services de Soins de Santé (ECPSS)* 2017. https://dhsprogram.com/publications/publication-SPA27-SPA-Final-Reports.cfm (2018).

24. Agence Nationale de la Statistique et de la Démographie (ANSD) & ICF. *Senegal: Enquête Continue sur la Prestation des Services de Soins de Santé (ECPSS)* 2019. https://dhsprogram.com/publications/publication-SPA33-SPA-Final-Reports.cfm (2020).

25. Ministère de la Santé et de l’’Action sociale (MSAS). *Plan National de Développement Sanitaire et Social (PNDSS) 2019-*2028. https://www.sante.gouv.sn/sites/default/files/1%20MSAS%20PNDSS%202019%202028%20Version%20Finale.pdf (2019).

26. Gueye, D. M. et al. A consolidated and geolocated list of health facilities in Senegal: consolidated list. XXXXX

27. Gueye, D. M. et al. A consolidated and geolocated list of health facilities in Senegal: full list. XXXXX

28. ESRI Sénégal. Carte sanitaire Kaolack. https://esrisenegal-esrisenegal.opendata.arcgis.com/datasets/carte-sanitaire-kaolack.

29. Jaro, M. A. Advances in Record-Linkage Methodology as Applied to Matching the 1985 Census of Tampa, Florida. Journal of the American Statistical Association 84, 414–420 (1989).

30. Robusto, C. C. The Cosine-Haversine Formula. The American Mathematical Monthly 64, 38–40 (1957).

31. Diop, I. L., Touré, I. D., Koita, M., Diop, M. & El-Khoury, M. *Cartographie du secteur privé de la santé au Sénégal 2016-*2017. https://shopsplusproject.org/sites/default/files/resources/Cartographie%20du%20secteur%20priv%C3%A9%20de%20la%20sant%C3%A9%20au%20S%C3%A9n%C3%A9gal%202016-2017_0.pdf (2018).

32. FERMETURE ANNONCEE DE L’HOPITAL NINEFECHA : VIVIANE WADE RECLAME SES CLEFS. SenePlus https://www.seneplus.com/article/fermeture-annoncee-de-l%E2%80%99hopital-ninefecha-viviane-wade-reclame-ses-clefs (2013).

33. Kédougou : L’hôpital de Ninéfécha transformé en poste de santé. Seneweb.com https://www.seneweb.com/news/Sante/kedougou-l-rsquo-hopital-de-ninefecha-tr_n_344829.html (2023).

34. Clinton Health Access Initiaitve (CHAI). Exemplars in geospatial data use for primary healthcare: cross-cutting lessons. https://clintonhealth.app.box.com/s/5ei753zgwz08njk8es6ttecq1xuuorim (2023).

35. Agence Nationale de la Statistique et de la Démographie (ANSD) & ICF. *Senegal: Enquête Démographique et de Santé Continue (EDS-Continue)* 2019. https://dhsprogram.com/publications/publication-FR368-DHS-Final-Reports.cfm (2020).

36. Africa CDC Annual Report 2021. Africa CDC https://africacdc.org/download/africa-cdc-annual-report-2021/.

37. Nguyen, M. et al. Mapping malaria seasonality in Madagascar using health facility data. BMC Medicine 18, 26 (2020).

38. Loquiha, O. et al. Mapping maternal mortality rate via spatial zero-inflated models for count data: A case study of facility-based maternal deaths from Mozambique. PLoS One 13, e0202186 (2018).

39. Croke, K., Mengistu, A. T., O’Connell, S. D. & Tafere, K. The impact of a health facility construction campaign on health service utilisation and outcomes: analysis of spatially linked survey and facility location data in Ethiopia. BMJ Global Health 5, e002430 (2020).

40. Gage, A. D., Fink, G., Ataguba, J. E. & Kruk, M. E. Hospital delivery and neonatal mortality in 37 countries in sub-Saharan Africa and South Asia: An ecological study. PLOS Medicine 18, e1003843 (2021).

41. Muhoza, P. et al. Key informant perspectives on the challenges and opportunities for using routine health data for decision-making in Senegal. BMC Health Services Research 21, 594 (2021).

42. Ministry of Health Malawi. Master Health Facility Registry of Malawi. https://zipatala.health.gov.mw/.

43. Federal Ministry of Health of Ethiopia. Master Facility Registry (MFR). https://mfrv2.moh.gov.et/#/dashboard.

44. R Core Team. R: A language and environment for statistical computing. (2021).

