## Supplementary information for "A consolidated and geolocated facility list in Senegal from triangulating secondary data"

**Table of contents** **Pages**


| Table of contents | Pages |
| --- | --- |
| Table S1. Total number of health facilities, by type and with geolocation, in Senegal and by region. | S2-3 |
| Table S2. Variable descriptions for the full facility list in Senegal. | S4-5 |
| Table S3. Comparing frequency of multiple facility types and facility ownership categorizations, by facility type, in Senegal and by region. | S6-7 |
| Table S4. Comparing GPS availability and concordance, by facility type, in Senegal and by region. | S8-9 |
| Table S5. Comparing number of facilities in the consolidated facility list with facility numbers reported in previously published facility lists by facility type, by region. | S10-11 |
| Table S6. Comparing number of facilities with GPS in the consolidated facility list with facility numbers from Maina and colleagues, nationally and by region. | S12 |
| Supplementary file 1. Report from the facility list workshop in Dakar, Senegal from January 31-February 1, 2023 (available in French only). *Please contact the corresponding author for access to this report.* | S13 |

**Table S1. Total number of health facilities, by type and with geolocation, in Senegal and by region.** All facilities here reflect hospitals, health centers, health posts, and health huts (i.e., not facilities designated as "other").

| Facility type | Location | Total facilities | Total facilities with GPS | Percentage of facilities with GPS (%) |
|---|---|---|---|---|
| **All facilities** | **Senegal** | **4,685** | **2,423** | **51.7%** |
| | Dakar | 797 | 328 | 41.2% |
| | Diourbel | 173 | 151 | 87.3% |
| | Fatick | 296 | 158 | 53.4% |
| | Kaffrine | 268 | 126 | 47.0% |
| | Kaolack | 370 | 218 | 58.9% |
| | Kedougou | 139 | 81 | 58.3% |
| | Kolda | 327 | 130 | 39.8% |
| | Louga | 582 | 191 | 32.8% |
| | Matam | 163 | 128 | 78.5% |
| | Saint-Louis | 265 | 204 | 77.0% |
| | Sedhiou | 204 | 102 | 50.0% |
| | Tambacounda | 317 | 161 | 50.8% |
| | Thies | 524 | 261 | 49.8% |
| | Ziguinchor | 260 | 184 | 70.8% |
| **Hospitals** | **Senegal** | **81** | **70** | **86.4%** |
| | Dakar | 35 | 30 | 85.7% |
| | Diourbel | 6 | 6 | 100% |
| | Fatick | 2 | 2 | 100% |
| | Kaffrine | 2 | 2 | 100% |
| | Kaolack | 4 | 1 | 25.0% |
| | Kedougou | 1 | 1 | 100% |
| | Kolda | 2 | 2 | 100% |
| | Louga | 3 | 2 | 66.7% |
| | Matam | 3 | 3 | 100% |
| | Saint-Louis | 5 | 5 | 100% |
| | Sedhiou | 3 | 2 | 66.7% |
| | Tambacounda | 1 | 1 | 100% |
| | Thies | 12 | 11 | 91.7% |
| | Ziguinchor | 2 | 2 | 100% |
| **Health centers** | **Senegal** | **257** | **227** | **88.3%** |
| | Dakar | 82 | 77 | 93.9% |
| | Diourbel | 11 | 8 | 72.7% |
| | Fatick | 9 | 9 | 100% |
| | Kaffrine | 6 | 4 | 66.7% |
| | Kaolack | 43 | 42 | 97.7% |
| | Kedougou | 5 | 5 | 100% |
| | Kolda | 9 | 7 | 77.8% |
| | Louga | 30 | 17 | 56.7% |
| | Matam | 5 | 5 | 100% |
| | Saint-Louis | 12 | 11 | 91.7% |
| | Sedhiou | 7 | 5 | 71.4% |

| Facility type | Location | Total facilities | Total facilities with GPS | Percentage of facilities with GPS (%) |
|---|---|---|---|---|
| | Tambacounda | 12 | 12 | 100% |
| | Thies | 16 | 15 | 93.8% |
| | Ziguinchor | 10 | 10 | 100% |
| **Health posts** | **Senegal** | **2,349** | **1,596** | **67.9%** |
| | Dakar | 680 | 221 | 32.5% |
| | Diourbel | 123 | 109 | 88.6% |
| | Fatick | 132 | 108 | 81.8% |
| | Kaffrine | 105 | 85 | 81.0% |
| | Kaolack | 151 | 132 | 87.4% |
| | Kedougou | 48 | 39 | 81.3% |
| | Kolda | 101 | 76 | 75.2% |
| | Louga | 161 | 126 | 78.3% |
| | Matam | 101 | 90 | 89.1% |
| | Saint-Louis | 136 | 125 | 91.9% |
| | Sedhiou | 75 | 61 | 81.3% |
| | Tambacounda | 162 | 118 | 72.8% |
| | Thies | 228 | 179 | 78.5% |
| | Ziguinchor | 146 | 127 | 87.0% |
| **Health huts** | **Senegal** | **1,998** | **530** | **26.5%** |
| | Dakar | - | - | - |
| | Diourbel | 33 | 28 | 84.8% |
| | Fatick | 153 | 39 | 25.5% |
| | Kaffrine | 155 | 35 | 22.6% |
| | Kaolack | 172 | 43 | 25.0% |
| | Kedougou | 85 | 36 | 42.4% |
| | Kolda | 215 | 45 | 20.9% |
| | Louga | 388 | 46 | 11.9% |
| | Matam | 54 | 30 | 55.6% |
| | Saint-Louis | 112 | 63 | 56.3% |
| | Sedhiou | 119 | 34 | 28.6% |
| | Tambacounda | 142 | 30 | 21.1% |
| | Thies | 268 | 56 | 20.9% |
| | Ziguinchor | 102 | 45 | 44.1% |

**Table S2. Variable descriptions for the full facility list in Senegal.** Variable descriptions for the full facility list are available in Table 4 of the manuscript.

| **Full facility list dataset** | |
|---|---|
| File name | senegal_full_facilitylist_full.csv |
| Total observations | 12,965 |
| **Full facility list variables** | |
| **Variable name** | **Variable description** |
| region | Region in Senegal. |
| department | Formal second-level administrative unit for Senegal, with n=45. *Note:* Data sources varied in their reporting of departments or health districts (and did not always specify which was included). Mapping capturing department information and accounting for inaccuracies occurred where possible, but some facility-source observations may not have a department included. |
| health_district | The peripheral level of Senegal's health sector, with an n=77 to 79. *Note:* Data sources varied in their reporting of departments or health districts (and did not always specify which was included). Mapping from district to department and accounting occurred where possible, but some facility-source observations may not have a health district included. |
| match_id | Facility id assigned for each unique facility grouping identified in the consolidated facility list. This identifier links facility groups in this dataset to unique observations found in the consolidated facility list. *Note:* this id does not correspond with a particular data source or formal health information system (e.g., DHIS2). |
| match_name | Processed version of the health facility name used for matching, excluding special characters, variations found across sources. |
| fac_name_orig | Original health facility name, as found in a given facility data source. |
| group_fac_type | This is the facility type assigned to a unique facility observation. Original facility types may vary by source, of which are listed under "fac_type_orig." Facility types included in this dataset are as follows in French (without special characters), with English translations: English translations:<br>- hopital= hospital<br>- centre de esante = health center<br>- poste de esante = health post<br>- case de sante= health hut<br>- autre = other |
| fac_type_orig | Original facility type, as provided in a given facility data source. |
| group_fac_own | Processed version of the managing authority for a given facility. Types included in this dataset are as follows in French (without special characters), with English translations:<br>- publique = public<br>- prive = private<br>- ong ou mission/confessionnel = NGO or mission/faith-based<br>- paramilitaire = paramilitary |
| fac_own_orig | Managing authority for the health facility, as provided in a given facility data source. |

| Full facility list variables | |
|---|---|
| **Variable name** | **Variable description** |
| latitude | Latitude coordinates, as provided in a given data source. |
| longitude | Longitude coordinates, as provided in a given data source. |
| max_gps_dist | Maximum Haversine distance in kilometers between GPS coordinates for each unique facility grouping. If a facility has only set of GPS coordinates (n_gps==1), this variable is blank. |
| source | Facility data source. |
| data_flagged | Variable whereby "1" indicates outstanding verification needs or follow-up questions. |
| data_notes | Further details about outstanding verification needs or follow-up questions about the given facility. All notes are in English at present. If there is information in "data_notes" but the facility is not flagged (data_flagged==1), these notes are meant to provide additional detail or information about a given facility without immediate action items or follow-up. |
| decision_notes | Further detail about decisions made for matching facilities and feedback from regional focal points. *All notes are in English at present.* |
| last_updated | Date of last update in YYYYMMDD format. For instance, 20230518 is 18 May 2023. These updates are meant to reflect changes to previous facility matches since the Dakar facility list workshop that occurred from 31 January to 1 February 2023. |
| update_notes | Further details on the updates that occurred for a unique facility observation. *All notes are in English at present.* |
| n_gps | Number of linked sources with GPS for each matched facility group.<br>*Note:* For a given facility, if multiple GPS coordinates were available and matched precisely, they were not counted as distinct sets of GPS. |
| n_source | Number of linked sources for each matched facility group. |
| source_list | List of sources for each matched facility group. |
| fac_id | Unique facility id, either directly provided from its original data source or generated in order to provide a unique identifier for each facility observation. |
| fac_id_orig | Original facility id, as provided in a given facility data source. *Note*: most non-survey data sources did not have an original facility id, and thus this variable is blank for them. |
| fac_id_orig_var | Variable name in which facility ids are contained, as found in a given facility data source.<br>*Note:* most non-survey data sources did not have an original facility id, and thus this variable is blank for them. |
| n_geos | Number of department-health district combinations within a matched facility group. This variable was included to more easily identify which matched facilities need further geographic variation, such that:<br>- If n_geos==0, then no department or health district information were included for these facilities.<br>- If n_geos==1, department-health district combinations for a matched facility group exist and are consistent.<br>- If n_geos >1, department-health district combinations vary within a matched facility group and further verification is needed. |

**Table S3. Comparing frequency of multiple facility types and facility ownership categorizations, by facility type, in Senegal and by region.** All facilities here reflect hospitals, health centers, health posts, and health huts. All facilities here reflect hospitals, health centers, health posts, and health huts (i.e., not facilities designated as "other").

| Facility type | Location | Total facilities | Total facilities with >1 facility ownership (% of total facilities) | Total facilities originally with >1 facility type (% of total facilities) | Total facilities originally with >1 facility type and then triangulated or verified (% of total facilities with > 1 facility type) |
|---|---|---|---|---|---|
| **All facilities** | **Senegal** | **4,685** | 1,895 (40.4%) | 486 (10.4%) | 202 (41.6%) |
| | Dakar | 797 | 232 (29.1%) | 95 (11.9%) | 48 (50.5%) |
| | Diourbel | 173 | 108 (62.4%) | 19 (11%) | 9 (47.4%) |
| | Fatick | 296 | 127 (42.9%) | 31 (10.5%) | 17 (54.8%) |
| | Kaffrine | 268 | 85 (31.7%) | 20 (7.5%) | 0 (0%) |
| | Kaolack | 370 | 132 (35.7%) | 42 (11.4%) | 27 (64.3%) |
| | Kedougou | 139 | 37 (26.6%) | 8 (5.8%) | 0 (0%) |
| | Kolda | 327 | 82 (25.1%) | 29 (8.9%) | 20 (69.0%) |
| | Louga | 582 | 239 (41.1%) | 34 (5.8%) | 5 (14.7%) |
| | Matam | 163 | 89 (54.6%) | 10 (6.1%) | 10 (100%) |
| | Saint-Louis | 265 | 139 (52.5%) | 44 (16.6%) | 9 (20.5%) |
| | Sedhiou | 204 | 112 (54.9%) | 17 (8.3%) | 7 (41.2%) |
| | Tambacounda | 317 | 198 (62.5%) | 82 (25.9%) | 20 (24.4%) |
| | Thies | 524 | 189 (36.1%) | 39 (7.4%) | 18 (46.2%) |
| | Ziguinchor | 260 | 126 (48.5%) | 16 (6.2%) | 12 (75.0%) |
| **Hospitals** | **Senegal** | **81** | 12 (34.3%) | 4 (11.4%) | 2 (50.0%) |
| | Dakar | 35 | 3 (50.0%) | 1 (16.7%) | 0 (0%) |
| | Diourbel | 6 | 2 (100%) | 2 (100%) | 1 (50.0%) |
| | Fatick | 2 | 1 (50.0%) | 1 (50.0%) | 0 (0%) |
| | Kaffrine | 2 | 1 (25.0%) | 1 (25.0%) | 0 (0%) |
| | Kaolack | 4 | 0 (0%) | 0 (0%) | - |
| | Kedougou | 1 | 2 (100%) | 0 (0%) | - |
| | Kolda | 2 | 0 (0%) | 0 (0%) | - |
| | Louga | 3 | 0 (0%) | 0 (0%) | - |
| | Matam | 3 | 1 (20.0%) | 1 (20.0%) | 0 (0%) |
| | Saint-Louis | 5 | 1 (33.3%) | 0 (0%) | - |
| | Sedhiou | 3 | 0 (0%) | 0 (0%) | - |
| | Tambacounda | 1 | 3 (25.0%) | 2 (16.7%) | 1 (50.0%) |
| | Thies | 12 | 0 (0%) | 1 (50.0%) | 0 (0%) |
| | Ziguinchor | 2 | 12 (34.3%) | 4 (11.4%) | 2 (50.0%) |
| **Health centers** | **Senegal** | **257** | 147 (57.2%) | 70 (27.2%) | 47 (67.1%) |
| | Dakar | 82 | 44 (53.7%) | 25 (30.5%) | 22 (88.0%) |
| | Diourbel | 11 | 8 (72.7%) | 3 (27.3%) | 3 (100%) |
| | Fatick | 9 | 8 (88.9%) | 5 (55.6%) | 3 (60.0%) |
| | Kaffrine | 6 | 5 (83.3%) | 2 (33.3%) | 0 (0%) |
| | Kaolack | 43 | 13 (30.2%) | 9 (20.9%) | 7 (77.8%) |
| | Kedougou | 5 | 4 (80.0%) | 0 (0%) | - |
| | Kolda | 9 | 9 (100%) | 3 (33.3%) | 1 (33.3%) |

| Facility type | Location | Total facilities | Total facilities with >1 facility ownership (% of total facilities) | Total facilities originally with >1 facility type (% of total facilities) | Total facilities originally with >1 facility type and then triangulated or verified (% of total facilities with > 1 facility type) |
|---|---|---|---|---|---|
| | Louga | 30 | 12 (40.0%) | 9 (30.0%) | 1 (11.1%) |
| | Matam | 5 | 4 (80.0%) | 2 (40.0%) | 2 (100%) |
| | Saint-Louis | 12 | 6 (50.0%) | 3 (25.0%) | 3 (100%) |
| | Sedhiou | 7 | 4 (57.1%) | 3 (42.9%) | 1 (33.3%) |
| | Tambacounda | 12 | 9 (75.0%) | 1 (8.3%) | 1 (100%) |
| | Thies | 16 | 11 (68.8%) | 3 (18.8%) | 1 (33.3%) |
| | Ziguinchor | 10 | 10 (100%) | 2 (20.0%) | 2 (100%) |
| **Health posts** | **Senegal** | **2,349** | 1,470 (62.6%) | 357 (15.2%) | 144 (40.3%) |
| | Dakar | 680 | 176 (25.9%) | 66 (9.7%) | 24 (36.4%) |
| | Diourbel | 123 | 97 (78.9%) | 15 (12.2%) | 6 (40.0%) |
| | Fatick | 132 | 113 (85.6%) | 23 (17.4%) | 13 (56.5%) |
| | Kaffrine | 105 | 77 (73.3%) | 17 (16.2%) | 0 (0%) |
| | Kaolack | 151 | 117 (77.5%) | 32 (21.2%) | 20 (62.5%) |
| | Kedougou | 48 | 33 (68.8%) | 7 (14.6%) | 0 (0%) |
| | Kolda | 101 | 68 (67.3%) | 18 (17.8%) | 13 (72.2%) |
| | Louga | 161 | 118 (73.3%) | 16 (9.9%) | 4 (25.0%) |
| | Matam | 101 | 85 (84.2%) | 8 (7.9%) | 8 (100%) |
| | Saint-Louis | 136 | 114 (83.8%) | 21 (15.4%) | 6 (28.6%) |
| | Sedhiou | 75 | 61 (81.3%) | 14 (18.7%) | 6 (42.9%) |
| | Tambacounda | 162 | 127 (78.4%) | 76 (46.9%) | 19 (25.0%) |
| | Thies | 228 | 171 (75.0%) | 31 (13.6%) | 15 (48.4%) |
| | Ziguinchor | 146 | 113 (77.4%) | 13 (8.9%) | 10 (76.9%) |
| **Health huts** | **Senegal** | 252 (12.6) | 46 (2.3%) | 7 (15.2%) | 252 (12.6%) |
| | Dakar | - | - | - | - |
| | Diourbel | 33 | 0 (0%) | 0 (0%) | - |
| | Fatick | 153 | 4 (2.6%) | 1 (0.7%) | 0 (0%) |
| | Kaffrine | 155 | 2 (1.3%) | 0 (0%) | - |
| | Kaolack | 172 | 1 (0.6%) | 0 (0%) | - |
| | Kedougou | 85 | 0 (0%) | 1 (1.2%) | 0 (0%) |
| | Kolda | 215 | 3 (1.4%) | 8 (3.7%) | 6 (75.0%) |
| | Louga | 388 | 109 (28.1%) | 9 (2.3%) | 0 (0%) |
| | Matam | 54 | 0 (0%) | 0 (0%) | - |
| | Saint-Louis | 112 | 18 (16.1%) | 19 (17.0%) | 0 (0%) |
| | Sedhiou | 119 | 46 (38.7%) | 0 (0%) | - |
| | Tambacounda | 142 | 62 (43.7%) | 5 (3.5%) | 0 (0%) |
| | Thies | 268 | 4 (1.5%) | 3 (1.1%) | 1 (33.3%) |
| | Ziguinchor | 102 | 3 (2.9%) | 0 (0%) | - |

**Table S4. Comparing GPS availability and concordance, by facility type, in Senegal and by region.** Facilities with more than 1 set of linked GPS coordinates and coordinates varying by more than 2 km could be considered of higher priority for further validation of their geolocation. For a given facility, if multiple GPS coordinates are available and they precisely match, they are not counted as distinct sets of GPS. All facilities here reflect hospitals, health centers, health posts, and health huts (i.e., not facilities designated as "other").

| Facility type | Location | Total facilities with GPS | Total facilities with > 1 set of GPS coordinates (% of total facilities with GPS) | Total facilities with > 1 set of GPS and > 2 km of each other (% of total facilities with > 1 set of GPS) |
|---|---|---|---|---|
| **All facilities** | **Senegal** | **2,423** | 1,433 (59.1%) | 384 (26.8%) |
| | Dakar | 328 | 171 (52.1%) | 28 (16.4%) |
| | Diourbel | 151 | 92 (60.9%) | 39 (42.4%) |
| | Fatick | 158 | 105 (66.5%) | 25 (23.8%) |
| | Kaffrine | 126 | 72 (57.1%) | 22 (30.6%) |
| | Kaolack | 218 | 120 (55.0%) | 27 (22.5%) |
| | Kedougou | 81 | 41 (50.6%) | 11 (26.8%) |
| | Kolda | 130 | 67 (51.5%) | 19 (28.4%) |
| | Louga | 191 | 111 (58.1%) | 31 (27.9%) |
| | Matam | 128 | 85 (66.4%) | 31 (36.5%) |
| | Saint-Louis | 204 | 133 (65.2%) | 37 (27.8%) |
| | Sedhiou | 102 | 66 (64.7%) | 19 (28.8%) |
| | Tambacounda | 161 | 92 (57.1%) | 26 (28.3%) |
| | Thies | 261 | 161 (61.7%) | 28 (17.4%) |
| | Ziguinchor | 184 | 117 (63.6%) | 41 (35.0%) |
| **Hospitals** | **Senegal** | **70** | 39 (55.7%) | 10 (25.6%) |
| | Dakar | 30 | 13 (43.3%) | 2 (15.4%) |
| | Diourbel | 6 | 4 (66.7%) | 0 (0%) |
| | Fatick | 2 | 2 (100%) | 1 (50.0%) |
| | Kaffrine | 2 | 1 (50.0%) | 0 (0%) |
| | Kaolack | 1 | 1 (100%) | 1 (100%) |
| | Kedougou | 1 | 0 (0%) | - |
| | Kolda | 2 | 2 (100%) | 1 (50.0%) |
| | Louga | 2 | 2 (100%) | 1 (50.0%) |
| | Matam | 3 | 2 (66.7%) | 1 (50.0%) |
| | Saint-Louis | 5 | 3 (60.0%) | 1 (33.3%) |
| | Sedhiou | 2 | 1 (50.0%) | 0 (0%) |
| | Tambacounda | 1 | 1 (100%) | 1 (100%) |
| | Thies | 11 | 5 (45.5%) | 1 (20.0%) |
| | Ziguinchor | 2 | 2 (100%) | 0 (0%) |
| **Health centers** | **Senegal** | **227** | 141 (62.1%) | 28 (19.9%) |
| | Dakar | 77 | 46 (59.7%) | 7 (15.2%) |
| | Diourbel | 8 | 6 (75.0%) | 2 (33.3%) |
| | Fatick | 9 | 8 (88.9%) | 0 (0%) |
| | Kaffrine | 4 | 4 (100%) | 2 (50.0%) |
| | Kaolack | 42 | 9 (21.4%) | 1 (11.1%) |
| | Kedougou | 5 | 4 (80.0%) | 1 (25.0%) |

| Facility type | Location | Total facilities with GPS | Total facilities with > 1 set of GPS coordinates (% of total facilities with GPS) | Total facilities with > 1 set of GPS and > 2 km of each other (% of total facilities with > 1 set of GPS) |
|---|---|---|---|---|
| | Kolda | 7 | 7 (100%) | 1 (14.3%) |
| | Louga | 17 | 11 (64.7%) | 3 (27.3%) |
| | Matam | 5 | 5 (100%) | 1 (20.0%) |
| | Saint-Louis | 11 | 6 (54.5%) | 3 (50.0%) |
| | Sedhiou | 5 | 5 (100%) | 2 (40.0%) |
| | Tambacounda | 12 | 9 (75.0%) | 2 (22.2%) |
| | Thies | 15 | 13 (86.7%) | 2 (15.4%) |
| | Ziguinchor | 10 | 8 (80.0%) | 1 (12.5%) |
| **Health posts** | **Senegal** | **1,596** | 1,185 (74.2%) | 320 (27.0%) |
| | Dakar | 221 | 112 (50.7%) | 19 (17.0%) |
| | Diourbel | 109 | 80 (73.4%) | 37 (46.2%) |
| | Fatick | 108 | 93 (86.1%) | 23 (24.7%) |
| | Kaffrine | 85 | 65 (76.5%) | 20 (30.8%) |
| | Kaolack | 132 | 109 (82.6%) | 25 (22.9%) |
| | Kedougou | 39 | 27 (69.2%) | 8 (29.6%) |
| | Kolda | 76 | 52 (68.4%) | 14 (26.9%) |
| | Louga | 126 | 93 (73.8%) | 24 (25.8%) |
| | Matam | 90 | 70 (77.8%) | 24 (34.3%) |
| | Saint-Louis | 125 | 108 (86.4%) | 27 (25.0%) |
| | Sedhiou | 61 | 50 (82.0%) | 13 (26.0%) |
| | Tambacounda | 118 | 80 (67.8%) | 22 (27.5%) |
| | Thies | 179 | 143 (79.9%) | 25 (17.5%) |
| | Ziguinchor | 127 | 103 (81.1%) | 39 (37.9%) |
| **Health huts** | **Senegal** | **530** | 68 (12.8%) | 26 (38.2%) |
| | Dakar | - | - | - |
| | Diourbel | 28 | 2 (7.1%) | 0 (0%) |
| | Fatick | 39 | 2 (5.1%) | 1 (50.0%) |
| | Kaffrine | 35 | 2 (5.7%) | 0 (0%) |
| | Kaolack | 43 | 1 (2.3%) | 0 (0%) |
| | Kedougou | 36 | 10 (27.8%) | 2 (20.0%) |
| | Kolda | 45 | 6 (13.3%) | 3 (50.0%) |
| | Louga | 46 | 5 (10.9%) | 3 (60.0%) |
| | Matam | 30 | 8 (26.7%) | 5 (62.5%) |
| | Saint-Louis | 63 | 16 (25.4%) | 6 (37.5%) |
| | Sedhiou | 34 | 10 (29.4%) | 4 (40.0%) |
| | Tambacounda | 30 | 2 (6.7%) | 1 (50.0%) |
| | Thies | 56 | 0 (0%) | - |
| | Ziguinchor | 45 | 2 (7.1%) | 0 (0%) |

**Table S5. Comparing number of facilities in the consolidated facility list with facility numbers reported in previously published facility lists by facility type, by region.** SPA 2012-2013 is omitted here as regional counts were not provided in the corresponding report. All facilities here reflect hospitals, health centers, health posts, and health huts (i.e., not facilities designated as "other").

| Region | Data source | All facilities | Hospitals | Health centers | Health posts | Health huts |
|---|---|---|---|---|---|---|
| **Dakar** | **Consolidated facility list** | **797** | **35** | **82** | **680** | **0** |
| | SPA 2017 | 721 | 35 | 39 | 647 | 0 |
| | SPA 2019 | | 42 | 41 | 664 | |
| | Health Map 2019 | | 16 | 107 | 638 | |
| | Health Map 2019 (public only) | | 15 | 23 | 129 | |
| | Health Map 2021 (public only) | 204 | 14 | 25 | 126 | 39 |
| **Diourbel** | **Consolidated facility list** | **173** | **6** | **11** | **123** | **33** |
| | SPA 2017 | 150 | 5 | 7 | 97 | 41 |
| | SPA 2019 | | 5 | 7 | 98 | |
| | Health Map 2019 | | 3 | 19 | 176 | |
| | Health Map 2019 (public only) | | 3 | 9 | 98 | |
| | Health Map 2021 (public only) | 234 | 4 | 9 | 108 | 113 |
| **Fatick** | **Consolidated facility list** | **296** | **2** | **9** | **132** | **153** |
| | SPA 2017 | 261 | 1 | 8 | 104 | 148 |
| | SPA 2019 | | 1 | 8 | 104 | |
| | Health Map 2019 | | 1 | 17 | 149 | |
| | Health Map 2019 (public only) | | 1 | 7 | 121 | |
| | Health Map 2021 (public only) | 220 | 1 | 4 | 98 | 117 |
| **Kaffrine** | **Consolidated facility list** | **268** | **2** | **6** | **105** | **155** |
| | SPA 2017 | 227 | 1 | 4 | 64 | 158 |
| | SPA 2019 | | 1 | 5 | 64 | |
| | Health Map 2019 | | 1 | 8 | 100 | |
| | Health Map 2019 (public only) | | 1 | 4 | 94 | |
| | Health Map 2021 (public only) | 375 | 1 | 4 | 116 | 254 |
| **Kaolack** | **Consolidated facility list** | **370** | **4** | **43** | **151** | **172** |
| | SPA 2017 | 296 | 4 | 6 | 97 | 189 |
| | SPA 2019 | | 8 | 6 | 98 | |
| | Health Map 2019 | | 1 | 13 | 183 | |
| | Health Map 2019 (public only) | | 1 | 4 | 113 | |
| | Health Map 2021 (public only) | 375 | 1 | 4 | 116 | 254 |
| **Kedougou** | **Consolidated facility list** | **139** | **1** | **5** | **48** | **85** |
| | SPA 2017 | 101 | 0 | 3 | 28 | 70 |
| | SPA 2019 | | 0 | 4 | 29 | |
| | Health Map 2019 | | | 6 | 47 | |
| | Health Map 2019 (public only) | | | 3 | 39 | |
| | Health Map 2021 (public only) | 130 | 1 | 4 | 42 | 83 |
| **Kolda** | **Consolidated facility list** | **327** | **2** | **9** | **101** | **215** |
| | SPA 2017 | 349 | 2 | 8 | 75 | 264 |
| | SPA 2019 | | 2 | 10 | 76 | |
| | Health Map 2019 | | 1 | 9 | 95 | |

| Region | Data source | All facilities | Hospitals | Health centers | Health posts | Health huts |
|---|---|---|---|---|---|---|
| | Health Map 2019 (public only) | | 1 | 4 | 69 | |
| | Health Map 2021 (public only) | 325 | 1 | 4 | 76 | 244 |
| **Louga** | **Consolidated facility list** | **582** | **3** | **30** | **161** | **388** |
| | SPA 2017 | 272 | 3 | 14 | 129 | 126 |
| | SPA 2019 | | 3 | 14 | 133 | |
| | Health Map 2019 | | 2 | 9 | 155 | |
| | Health Map 2019 (public only) | | 2 | 4 | 116 | |
| | Health Map 2021 (public only) | 480 | 2 | 10 | 118 | 350 |
| **Matam** | **Consolidated facility list** | **163** | **3** | **5** | **101** | **54** |
| | SPA 2017 | 136 | 1 | 5 | 76 | 54 |
| | SPA 2019 | | 1 | 5 | 76 | |
| | Health Map 2019 | | 2 | 9 | 106 | |
| | Health Map 2019 (public only) | | 2 | 4 | 96 | |
| | Health Map 2021 (public only) | 186 | 3 | 9 | 102 | 72 |
| **Saint-Louis** | **Consolidated facility list** | **265** | **5** | **12** | **136** | **112** |
| | SPA 2017 | 214 | 2 | 6 | 111 | 95 |
| | SPA 2019 | | 2 | 6 | 111 | |
| | Health Map 2019 | | 3 | 21 | 154 | |
| | Health Map 2019 (public only) | | 3 | 8 | 112 | |
| | Health Map 2021 (public only) | 321 | 3 | 9 | 123 | 186 |
| **Sedhiou** | **Consolidated facility list** | **204** | **3** | **7** | **75** | **119** |
| | SPA 2017 | 118 | 1 | 4 | 43 | 70 |
| | SPA 2019 | | 1 | 4 | 43 | |
| | Health Map 2019 | | 1 | 6 | 72 | |
| | Health Map 2019 (public only) | | 1 | 3 | 62 | |
| | Health Map 2021 (public only) | 167 | 1 | 4 | 60 | 102 |
| **Tambacounda** | **Consolidated facility list** | **317** | **1** | **12** | **162** | **142** |
| | SPA 2017 | 193 | 1 | 18 | 79 | 95 |
| | SPA 2019 | | 1 | 17 | 59 | |
| | Health Map 2019 | | 1 | 20 | 155 | |
| | Health Map 2019 (public only) | | 1 | 7 | 125 | |
| | Health Map 2021 (public only) | 284 | 1 | 7 | 148 | 128 |
| **Thies** | **Consolidated facility list** | **524** | **12** | **16** | **228** | **268** |
| | SPA 2017 | 486 | 10 | 14 | 177 | 285 |
| | SPA 2019 | | 11 | 14 | 178 | |
| | Health Map 2019 | | 5 | 24 | 349 | |
| | Health Map 2019 (public only) | | 3 | 9 | 180 | |
| | Health Map 2021 (public only) | 492 | 5 | 10 | 180 | 297 |
| **Zinguichor** | **Consolidated facility list** | **260** | **2** | **10** | **146** | **102** |
| | SPA 2017 | 240 | 2 | 12 | 126 | 100 |
| | SPA 2019 | | 2 | 12 | 126 | |
| | Health Map 2019 | | 2 | 14 | 184 | |
| | Health Map 2019 (public only) | | 2 | 5 | 124 | |
| | Health Map 2021 (public only) | 237 | 2 | 5 | 114 | 116 |

**Table S6. Comparing number of facilities with GPS in the consolidated facility list with facility numbers from Maina and colleagues, nationally and by region.** All facilities here reflect hospitals, health centers, health posts, and health huts (i.e., not facilities designated as "other").

| Region | Data source | All facilities | Hospitals | Health centers | Health posts | Health huts |
|---|---|---|---|---|---|---|
| **Senegal** | **Consolidated facility list** | **2,423** | **70** | **227** | **1,596** | **530** |
| | Maina et al 2019 | 1,256 | 29 | 83 | 1,144 | |
| **Dakar** | **Consolidated facility list** | **328** | **30** | **77** | **221** | |
| | Maina et al 2019 | 121 | 10 | 14 | 97 | |
| **Diourbel** | **Consolidated facility list** | **151** | **6** | **8** | **109** | **28** |
| | Maina et al 2019 | 91 | 3 | 6 | 82 | |
| **Fatick** | **Consolidated facility list** | **158** | **2** | **9** | **108** | **39** |
| | Maina et al 2019 | 99 | 1 | 6 | 92 | |
| **Kaffrine** | **Consolidated facility list** | **126** | **2** | **4** | **85** | **35** |
| | Maina et al 2019 | 71 | 1 | 4 | 66 | |
| **Kaolack** | **Consolidated facility list** | **218** | **1** | **42** | **132** | **43** |
| | Maina et al 2019 | 77 | 1 | 2 | 74 | |
| **Kedougou** | **Consolidated facility list** | **81** | **1** | **5** | **39** | **36** |
| | Maina et al 2019 | 30 | | 3 | 27 | |
| **Kolda** | **Consolidated facility list** | **130** | **2** | **7** | **76** | **45** |
| | Maina et al 2019 | 53 | 1 | 3 | 49 | |
| **Louga** | **Consolidated facility list** | **191** | **2** | **17** | **126** | **46** |
| | Maina et al 2019 | 113 | 2 | 12 | 99 | |
| **Matam** | **Consolidated facility list** | **128** | **3** | **5** | **90** | **30** |
| | Maina et al 2019 | 79 | 2 | 4 | 73 | |
| **Saint-Louis** | **Consolidated facility list** | **204** | **5** | **11** | **125** | **63** |
| | Maina et al 2019 | 119 | 2 | 4 | 113 | |
| **Sedhiou** | **Consolidated facility list** | **102** | **2** | **5** | **61** | **34** |
| | Maina et al 2019 | 49 | 1 | 2 | 46 | |
| **Tambacounda** | **Consolidated facility list** | **161** | **1** | **12** | **118** | **30** |
| | Maina et al 2019 | 88 | 1 | 7 | 80 | |
| **Thies** | **Consolidated facility list** | **261** | **11** | **15** | **179** | **56** |
| | Maina et al 2019 | 156 | 2 | 11 | 143 | |
| **Zinguichor** | **Consolidated facility list** | **184** | **2** | **10** | **127** | **45** |
| | Maina et al 2019 | 110 | 2 | 5 | 103 | |

**Supplementary file 1. Report from the facility list workshop in Dakar, Senegal from January 31-February 1, 2023 (available in French only).** *Please contact the corresponding author for access to this report.*
